# Transcriptomic pathology of neocortical microcircuit cell types across psychiatric disorders

**DOI:** 10.1101/2023.10.26.23297640

**Authors:** Keon Arbabi, Dwight F. Newton, Hyunjung Oh, Melanie C. Davie, David A. Lewis, Michael Wainberg, Shreejoy J. Tripathy, Etienne Sibille

## Abstract

**Background:** Psychiatric disorders like major depressive disorder (MDD), bipolar disorder (BD), and schizophrenia (SCZ) are characterized by altered cognition and mood, brain functions that depend on information processing by cortical microcircuits. We hypothesized that psychiatric disorders would display cell type-specific transcriptional alterations in neuronal subpopulations that make up cortical microcircuits: excitatory pyramidal (PYR) neurons and vasoactive intestinal peptide- (VIP), somatostatin- (SST), and parvalbumin- (PVALB) expressing inhibitory interneurons.

**Methods:** We performed cell type-specific molecular profiling of subgenual anterior cingulate cortex, a region implicated in mood and cognitive control, using laser capture microdissection followed by RNA sequencing (LCM-seq). We sequenced libraries from 130 whole cells pooled per neuronal subtype (VIP, SST, PVALB, superficial and deep PYR) in 76 subjects from the University of Pittsburgh Brain Tissue Donation Program, evenly split between MDD, BD, and SCZ subjects and healthy controls.

**Results:** We identified hundreds of differentially expressed (DE) genes and biological pathways across disorders and neuronal subtypes, with the vast majority in inhibitory neuron types, primarily PVALB. DE genes were distinct across cell types, but partially shared across disorders, with nearly all shared genes involved in the formation and maintenance of neuronal circuits. Coordinated alterations in biological pathways were observed between select pairs of microcircuit cell types and partially shared across disorders. Finally, DE genes coincided with known risk variants from psychiatric genome-wide association studies, indicating cell type-specific convergence between genetic and transcriptomic risk for psychiatric disorders.

**Conclusions:** We present the first cell type-specific dataset of cortical microcircuit gene expression across multiple psychiatric disorders. Each neuronal subtype displayed unique dysregulation signatures, some shared across cell types and disorders. Inhibitory interneurons showed more dysregulation than excitatory pyramidal neurons. Our study suggests transdiagnostic cortical microcircuit pathology in SCZ, BD, and MDD and sets the stage for larger-scale studies investigating how cell circuit-based changes contribute to shared psychiatric risk.

## Introduction

Psychiatric disorders like major depressive disorder (MDD), bipolar disorder (BD), and schizophrenia (SCZ) are associated with widespread molecular alterations in the brain, including alterations in neurotransmitter systems (serotonin, dopamine, norepinephrine, glutamate, GABA) (1–7), deficits in neuronal homeostasis (8,9), neuroinflammation (10–12), and oxidative and endoplasmic reticulum stress (13–17). They are also associated with brain circuit dysfunction at multiple scales, including large-scale brain networks like the default mode network (18,19), excitatory-inhibitory balance (20–22), and cortical microcircuits (23–27).

Cortical microcircuits are modules of brain cells connected in an evolutionarily conserved pattern, consisting of a single excitatory (glutamatergic) pyramidal neuron that feeds into three primary inhibitory (GABAergic) interneuron types: parvalbumin-expressing (PVALB), somatostatin-expressing (SST), and vasointestinal peptide-expressing (VIP). These interneuron types provide feedback back onto the pyramidal neuron, completing the circuit (**Figure 1A**). This interconnected structure constitutes the fundamental information processing unit of cortical and hippocampal areas (28). Molecular changes in any of the neuronal types that make up cortical microcircuit can disrupt cellular function and neural information processing. It is thought that these lower-level alterations are in large part responsible for the higher-level circuit alterations observed in psychiatric disorders, including excitatory-inhibitory imbalance and alterations in large-scale brain networks, which in turn contribute to psychiatric symptoms (26).

**Figure 1:**
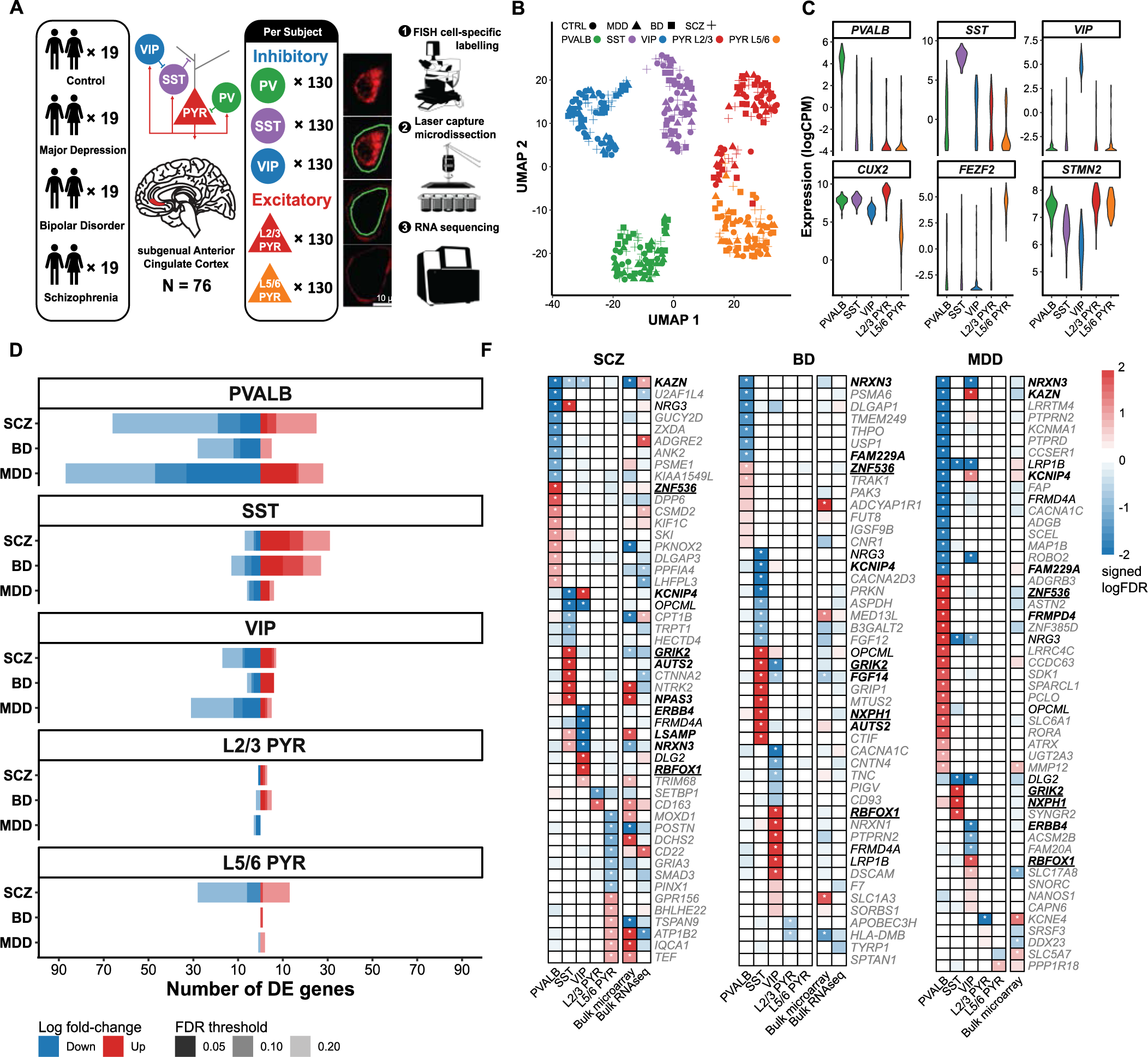
LCM-seq and differential expression testing identify cell type-specific transcriptomic perturbations in interneurons that are shared in part across psychiatric disorders. **A)** Overview of experimental strategy. LCM-seq was used to survey cortical microcircuit neuronal subtypes in sgACC samples from 19 tetrads of control, major depressive disorder (MDD), bipolar disorder (BD), and schizophrenia (SCZ) subjects matched on age, sex, and tissue quality (N = 76 subjects total). **B)** UMAP plot of gene expression (top 2000 variable features) shows separation of L2/3 PYR, L5/6 PYR, PVALB, VIP and SST cells from each subject. **C)** Normalized expression of cell type-specific (*PVALB, SST, VIP, CUX2 = L2/3 PYR, FEZF2 = L5/6 PYR*) and general neuronal (*STMN2*) marker genes. **D)** Number of up-regulated (red) and down-regulated (blue) differentially expressed (DE) genes at varying FDR thresholds (0.05, 0.10, 0.20). **E)** Top DE genes for each cell type and disorder by significance level, and comparison prior meta-analysis of microarray and RNA-seq gene expression in bulk neocortical samples (45) (signed −log10 FDR, * = FDR < 0.20). Black colored genes are DE in the same cell type for at least two disorders but have opposite fold change signs. Bolded genes and bolded underlined genes are DE in the same direction in the same cell type for two or all three disorders, respectively (not all shared genes shown, see Supplemental Figure 5).

Cell type-specific molecular changes in psychiatric disorders have been most widely studied through gene expression, primarily using two technologies: single-nucleus RNA sequencing (snRNA-seq) and laser capture microdissection followed by RNA sequencing (LCM-seq). snRNA-seq, in which each RNA molecule is tagged with its cell of origin, has been performed on postmortem brain samples from patients with MDD (29,30), MDD and PTSD (31), and SCZ (32–34). snRNA-seq offers the highest throughput, but with the disadvantages of only capturing RNA from inside the nucleus and of lower transcript coverage. LCM-seq uses lasers to cut out (microdissect) entire neuron bodies – not just the nucleus – that are then pooled by cell type for RNA sequencing, and has enabled whole-cell characterization of pyramidal neurons in SCZ and schizoaffective disorder (35), pyramidal neurons in BD and MDD (36), PVALB neurons in SCZ (37), and stellate neurons in SCZ (38). LCM-seq is especially well-suited for studying cortical microcircuits, as only neuronal cell types of interest are sequenced and coverage of relatively rare cell types like neocortical interneurons is enhanced, enabling balanced sampling of cells and reads across cell types and subjects. To date, no study has examined whole-cell gene expression changes across distinct neuronal subtypes forming functional circuits and across psychiatric disorders simultaneously.

In this study, we generated the first cell type-specific dataset of cortical microcircuit gene expression across multiple psychiatric disorders. We performed LCM-seq on the postmortem subgenual anterior cingulate cortex of 76 subjects, evenly split between MDD, BD, and SCZ cases and healthy controls (19 subjects per group). We performed differential expression testing for each disorder and neuronal subtype, both at the level of individual genes and pathways, and identified shared alterations across disorders that converge on risk loci from psychiatric genome-wide association studies.

## Methods

### Human subjects

We obtained brain samples during autopsies conducted at the Allegheny County Medical Examiner’s Office in Pittsburgh, PA, after we procured consent from the next of kin. An independent committee of clinical research scientists at the University of Pittsburgh made consensus DSM-IV diagnoses using clinical records, toxicology results, and standardized psychological autopsies. We included 76 subjects in our study, divided into 19 groups of 4 subjects (“tetrads”): one with major depressive disorder (MDD), one with bipolar disorder (BD), one with schizophrenia (SCZ), and one without any psychiatric disorder (control). The four subjects in each tetrad were matched for sex and as closely as possible for age. Groups did not differ in mean postmortem interval (PMI), pH, or RIN. We sectioned fresh-frozen tissue blocks containing the subgenual anterior cingulate cortex (sgACC) at 12 μm thickness, mounted these on RNase-free laser capture microdissection (LCM)-compatible slides, and stored them at −80°C until we harvested the cells. All procedures were approved by the University of Pittsburgh Committee for Oversight of Research and Clinical Training Involving Decedents and Institutional Review Board for Biomedical Research. We describe the demographic and clinical characteristics of our cohort in **Supplemental Table 1**.

### Cell density analysis

We performed double-label fluorescent *in situ* hybridization (FISH) on sgACC sections, targeting *Slc17a7* (a marker of pyramidal cells) + *Sst*, or *Pvalb* + *Vip*, counterstained with DAPI to determine changes in cell type density. We performed RNAscope staining in accordance with the manufacturer’s instructions. We adopted a stereological approach to randomly sample 20 sites of the sgACC for imaging with an Olympus IX3 confocal microscope (Olympus, Tokyo, Japan). We distinguished L2/3 and L5/6 pyramidal (PYR) cells through the selection of sites superficial and deep to a reference line midway between the pial surface and white matter. We obtained Z-stacks of images that represented transverse depth into tissue sections at a step size of 0.25 μm. We imaged each site to a depth of 12 Z-stacks, reflecting the two boundaries of the tissue beyond which our microscope focus could not be achieved. We deconvolved images using the AutoQuant Blind algorithm in Slidebook version 6.0 (Intelligent Imaging Innovations, Denver, CO) to reduce background; we collapsed Z-stacks into a composite image, normalizing intensities across all samples. We manually quantified cells containing 10 or more cell-marker grains and overlapping with DAPI. We adjusted cell type densities for age, sex, and PMI (**Supplemental Figure 1**).

### Fluorescent labeling and capture of microcircuit neuronal subtypes

We used a rapid FISH protocol, a shortened version of the RNAscope Multiplex Fluorescent V2 kit protocol (Advanced Cell Diagnostics, Newark, CA), to stain sgACC sections with cDNA probes specific to *Slc17a7*, *Sst*, *Pvalb*, or *Vip*, thereby visualizing microcircuit cell types, as previously described (39,40). Just as in our cell density analyses, we identified L2/3 and L5/6 PYR-cells using *Slc17a7* and distinguished them with a reference line placed halfway between the pial surface and white matter. We used one probe per tissue section and made all probes visible with the Opal 520 dye (FITC range) to minimize detection bias across cell types. We counter-stained sections with Neurotrace Red (ThermoFisher, Waltham, MA), a fluorescent Nissl stain, to visualize cell bodies. We then collected 130 of each cell type from each subject using an LMD7 laser microdissection microscope (Leica Microsystems). Altogether, we dissected and captured 49,400 neurons across all samples.

### Library preparation and RNA-sequencing

We extracted RNA using a PicoPure RNA isolation kit (Thermo Fisher Scientific, Waltham, MA), and prepared libraries with the SMARTer Stranded Total RNA-Seq Pico Input Kit V2 (Clontech, Mountain View, CA). We determined library fragment size distribution with a Bioanalyzer (Agilent Technologies, Santa Clara, CA), and carried out initial quality control sequencing on an Illumina MiSeq sequencer (Illumina, San Diego, CA). Given the limited RNA quantity from each pool of our microdissected neurons, we did not assess RNA integrity number (RIN) values in the neuron samples used for RNA-seq profiling. However, we have previously shown that our Nissl staining and laser microdissection method results in RIN >7 in all samples, and values are nearly identical to those obtained in tissue homogenates from the same subjects (35). If libraries showed < 80% reads aligning to the genome, or < 15% of reads aligning to exons, we re-collected them (i.e., repeated the RNAscope-LCM procedure using new tissue sections) and sequenced again. We loaded 3 or 4 tetrads on individual flow cells, using 6 flow-cells in total: 5 for initial sequencing and 1 for re-collected cells. Libraries that passed quality control (N = 379/380, 1 subject had insufficient gray matter for PV-cell collection) were sequenced on an Illumina NovaSeq sequencer (Illumina, San Diego, CA) to generate 2 x 125 bp paired-end reads. We pooled 3 or 4 tetrads and loaded onto each flow cell, using 5 NovaSeq S2 flow-cells.

We aligned paired-end reads to GRCh38 using the Ensembl annotation (Release 98) with alignment performed using STAR v2.7.2 with default parameters (41). We used the summarizeOverlaps function from the GenomicAlignments R package to quantify genes, with separate quantifications for exon- and intron-aligning reads (42). Before gene quantification, we utilized the ScanBamParam function from Rsamtools to extract read information, exclude PCR/optical duplicates, and filter for uniquely aligning reads that mapped to their proper read pair. NovaSeq sequencing yielded an average of 48,457,414 reads per cell type per subject (SE ± 377,012), with an average genomic mapping rate of 78.39% (SE ± 0.41%; 38,124,400 reads ± 388,752), exonic alignment rate of 31.48% (SE ± 0.39%), and intronic alignment rate of 49.26% (SE ± 0.15%). We combined exonic and intronic reads and removed mitochondrially encoded genes (genes with the *MT-* prefix) for downstream analysis steps.

### Differential expression analysis

We performed gene filtering and differential expression (DE) analysis separately for each neuronal cell type. We included genes with > 1 count in ≥ 80% of biological replicates. After normalizing counts and calculating precision weights using voom, we computed the variance of gene expression using linear models implemented in the limma package (43). The main independent variable for our models was the diagnostic group (MDD, BD, SCZ versus control), and we also adjusted for sex and intergenic rate.

In addition to diagnostic group, we considered whether to include each of 18 biological and technical covariates in our models. Biological covariates comprised sex, age, race, method of death (MOD), tobacco use, medication use at time of death (antipsychotics, antidepressants, lithium, benzodiazepines and/or anticonvulsants), while technical covariates comprised pH, RIN, PMI, pool (batch), number of detected genes, library size, intronic rate, intergenic rate, and intragenic rate. Many of these covariates were highly correlated with each other (**Supplemental Figure 2A**) and were correlated with principal components (PCs) of the normalized gene expression data (**Supplemental Figure 2B**). We selected the final covariates empirically within each cell type using the Bayesian Information Criterion (BIC) (44) with the mvForwardStepwise function from the mvIC R package. One phase of covariate selection was performed on biological covariates, and a second phase, including significant covariates from the first phase, was performed on technical covariates. Of the 18 covariates, the ones selected were sex (all five cell types), intergenic rate (all cell types except L2/3 PYR), race (PVALB and L5/6 PYR only), and MOD (L2/3 PYR and L5/6 PYR only). We included only sex and intergenic rate in the final linear models to ensure a consistent interpretation of DE across cell types. Importantly, medication use covariates were not significantly correlated with gene expression PCs or selected by mvForwardStepwise.

We considered genes with a Benjamini-Hochberg adjusted p-value (FDR) < 0.20 to be DE. This exploratory threshold enabled us to assess the overlap between cell types and disorders. We also compared our DE genes to those from a high-quality meta-analysis of bulk cortex microarray and RNA-seq data from cerebral cortex (45). Additionally, we evaluated the specificity of cell type-specific DE genes using an external single nucleus RNA-seq dataset of human medial temporal gyrus from the Allen Institute (46). First, we mapped each of the five LCM-seq neuronal subtypes to its closest snRNA-seq ‘subclass’ based on maximal expression of subclass-specific marker genes. We considered DE genes cell type-specific if at least 10% of cells of the respective snRNA-seq subclass expressed that gene (**Supplemental Figure 3B**). We also calculated the gene-wise correlation of expression levels between LCM-seq controls and the snRNA-seq reference to assess concordance between these sequencing methods (**Supplemental Figure 3C**).

We further compared the concordance between LCM-seq gene signatures and recent snRNA-seq datasets of MDD and SCZ (30,34). We obtained snRNA-seq count matrices, filtered out low-quality cells and genes, and performed pseudobulk DE testing using limma-voom (43). LCM-seq cell types were manually matched to the authors’ original cell type annotations based on semantic similarities (e.g., InN9_PV mapped to PVALB) and correlation of DE t-statistics across genes. To assess the overlap of gene signatures between LCM-seq and snRNA-seq in specific cell types, we employed the rank-rank hypergeometric overlap test (RRHO) (47). Genes were ranked according to their p-value and effect size direction, allowing us to identify significantly overlapping genes across a continuous range of significance levels (**Supplemental Figure 4**).

### Differential pathway activity analysis

We curated disease-associated pathways (gene sets) by filtering Gene Ontology (GO) biological processes (downloaded from the Mayaan laboratory; https://maayanlab.cloud/Enrichr) to pathways whose names contained manually curated keywords related to biological processes known to be affected in brain disorders, including bioenergetics, metabolism, transcription and translation, growth factors, cytoskeleton, cellular trafficking, neuron structure and function, and immune system and inflammation (see **Supplemental Table 2** for the list of keywords). We manually curated lists to ensure the alignment of the pathways with the chosen biological themes. We performed differential pathway activity analysis similarly to methods published previously (48). We used Gene Set Variation Analysis (GSVA) (49) to summarize normalized gene expression into activity scores for each pathway, set to the following parameters: mx.diff = TRUE, kcdf = “Gaussian”, min.sz = 5, max.sz = 150. Briefly, GSVA calculates a normalized relative expression level per gene across samples. This expression level is then rank-ordered for each sample and aggregated into a pathway-level activity score by calculating sample-wise enrichment scores using a Kolmogorov–Smirnov-like rank statistic. To minimize false positives, we applied certain filters: genes that were not expressed in a given cell type (<1 CPM in ≥70% of samples) were excluded, as well as pathways for which >30% of the genes in the set were not expressed. For each cell type and pathway, we modeled pathway activity as a function of diagnosis, sex, and intergenic rate using lmFit in limma (43). Pathways demonstrating evidence of association with diagnosis at p < 0.01 in at least one cell type and disorder were included in the final analyses.

Next, we identified coordinated alterations in biological pathways within the cortical microcircuit associated with each disorder. To achieve this, we examined the DE *t*-statistic of all genes defining the pathways within each biological theme and correlated them across cell types pairs. A significant (p < 0.05) positive Pearson correlation between two cell types was interpreted as a coordinated biological change for that disease.

### Overlap with genetic risk loci

We evaluated the intersection of cell type-specific DE genes and psychiatric disorder-associated risk genes using PoPS and PsyOPS, advanced gene-prioritization methods for GWAS (61,62). PoPS integrates multiple gene features, such as cell-type-specific expression and biological pathways, using aggregated gene-level p-values from GWAS summary statistics. PsyOPS is trained on three features that hypothesize causal genes are mutationally constrained, brain-specific, and overlap with known neurodevelopmental genes. For this analysis, we provided both tools with GWAS summary statistics from recent large-scale studies on MDD, BD, and SCZ (50–52). After running the tools as prescribed, we merged their outputs with our DE gene list (FDR < 0.20). The final dataset included only GWAS loci that were highlighted by at least one tool and overlapped with a disorder-specific DE gene (**Figure 3B**).

## Results

### LCM-seq generates robust cell type-specific transcriptomes from cortical microcircuit neuronal subtypes in postmortem human brain tissues

We performed laser capture microdissection (LCM) followed by RNA sequencing (LCM-seq) on postmortem samples from the subgenual anterior cingulate cortex (sgACC) of 76 subjects. These subjects comprised 19 groups of 4 subjects (“tetrads”) matched closely on age, sex, and tissue quality: one with major depressive disorder (MDD), one with bipolar disorder (BD), one with schizophrenia (SCZ), and one with no psychiatric disorder (control; **Figure 1A**, **Table 1**). We achieved cell type specificity using RNAscope-based FISH probes to identify each of five neuronal subtypes: Layer 2/3 and layer 5/6 glutamatergic pyramidal neurons (L2/3, L5/6 PYR), as well as vasoactive intestinal peptide- (VIP), somatostatin- (SST), and parvalbumin- (PVALB) expressing inhibitory interneurons. We then used LCM to collect 130 cells for each neuron type per subject prior to performing RNA sequencing. Thus, we generated 76 (subjects) × 5 (cell types) = 380 bulk transcriptomes, each containing the pooled RNA from 130 cells.

**Table 1.** Subject characteristics. Mean ± SD are shown for continuous variables and count (percentage) for categorical variables. Significance of diagnostic group differences is assessed via ANOVA p-value (for categorical variables) or chi-squared (for continuous); significant p-values are bolded. Abbreviations: AUD = Alcohol use disorder, SUD = Substance use disorder, ATOD = At time of death.

|  | Control<br>(N = 19) | MDD<br>(N = 19) | BD<br>(N = 19) | SCZ<br>(N = 19) | Group Differences p-value<br>(ANOVA or chi-squared) |
| --- | --- | --- | --- | --- | --- |
| Age (Years) | 46.3 ± 9.5 | 47.8 ± 10.4 | 45.2 ± 10.1 | 45.1 ± 8.5 | 0.900 |
| Sex (Female) | 9 (47%) | 9 (47%) | 9 (47%) | 9 (47%) | 1.0 |
| Race (White) | 18 (95%) | 18 (95%) | 18 (95%) | 13 (68%) | **0.012** |
| PMI (Hours) | 21.3 ± 6.6 | 19.3 ± 5.3 | 20.1 ± 6 | 20.1 ± 6.9 | 0.819 |
| pH | 6.6 ± 0.2 | 6.6 ± 0.2 | 6.6 ± 0.2 | 6.6 ± 0.3 | 0.935 |
| RIN | 8.0 ± 0.4 | 8.0 ± 0.6 | 8.0 ± 0.5 | 7.9 ± 0.7 | 0.902 |
| Suicide | 0 (0%) | 7 (37%) | 8 (42%) | 7 (37%) | 1.0 |
| AUD ATOD | 0 (0%) | 3 (16%) | 4 (21%) | 5 (26%) | 0.918 |
| SUD ATOD | 0 (0%) | 4 (21%) | 6 (32%) | 3 (16%) | 0.625 |
| Either AUD/SUD ATOD | 0 (0%) | 6 (32%) | 8 (42%) | 6 (32%) | 0.829 |
| Tobacco use ATOD | 6 (26%) | 7 (37%) | 9 (47%) | 12 (63%) | **0.033** |
| Benzodiazepines | 0 (0%) | 3 (16%) | 9 (42%) | 4 (21%) | 0.084 |
| Anticonvulsants | 0 (0%) | 1 (5%) | 7 (37%) | 6 (32%) | 0.057 |
| Antidepressants | 0 (0%) | 7 (37%) | 12 (63%) | 10 (53%) | 0.306 |
| Lithium | 0 (0%) | 0 (0%) | 2 (11%) | 0 (0%) | 0.321 |
| Antipsychotic | 0 (0%) | 2 (11%) | 7 (37%) | 16 (84%) | **4.0 × 10<sup>-6</sup>** |
| Any Medication | 0 (0%) | 8 (42%) | 15 (79%) | 16 (84%) | **0.008** |

In UMAP space, we observed a clustering of transcriptomes primarily by cell type (93.8% accuracy, k-means clustering, k = 5), with some minor overlap between L2/3 and L5/6 PYR cells (**Figure 1B**). As expected, each cell type had enriched expression of cell type-specific marker genes (*PVALB, SST, VIP, CUX2 =* L2/3 PYR*, FEZF2 =* L5/6 PYR, all p < 1.0 × 10^−4^) (**Figure 1C**). Further demonstrating the cell type-specificity of the data, gene expression levels from LCM-seq profiled cell types exhibited a strong correlation with those from a single-nucleus human neocortical RNA-seq reference atlas (46) (r = 0.85 ± 0.0021, all p < 2.6 × 10^−16^) (**Supplemental Figure 3B**). In parallel tissue sections from those used for cell type collection, we determined cell densities using FISH. No cell types showed significant differences in microscopy-based cellular density between cases and controls after adjusting for sex, age and PMI, although trending decreases in interneuron densities were observed for SCZ (**Supplemental Figure 1**).

### Differential expression analysis highlights distinct transcriptional alterations in inhibitory interneuron subtypes, partially shared across psychiatric disorders

To identify disease-associated transcriptional alterations, we performed case-control differential expression (DE) testing separately for each cell type. A total of 460 DE genes (N = 371 unique) were identified across cell types and disorders at FDR < 0.20, with the majority (N = 398, 87.3%) observed in inhibitory interneurons (**Figure 1D**). PVALB cells displayed the greatest number of DE genes (N = 239), with 91 for SCZ (25 upregulated, 66 downregulated), 33 for BD (5 up, 28 down), and 113 for MDD (28 up, 85 down). For SST cells, there were 38 DE genes for SCZ (31 up, 7 down), 40 for BD (27 up, 13 down), and 12 for MDD (6 up, 6 down). For VIP cells, there were 24 DE genes for SCZ (7 up, 17 down), 11 for BD (6 up, 5 down), and 36 for MDD (5 up, 31 down). The majority of DE in excitatory neurons was observed in L5/6 PYR for SCZ (N = 41, 13 up, 28 down). See **Supplemental Table 1** for the complete DE analysis results.

Figure 1E presents the top altered genes across neuronal subtypes in each disorder, ordered by significance (FDR). Although, the majority of DE genes (N = 314, 84.6%) were perturbed in a single neuronal subtype, 9.2% (N = 34) of DE genes were altered in the same direction and cell type across at least two disorders (p = 2.6 × 10^−33^, hypergeometric test). In other words, disease-associated expression changes were highly cell type-specific; however, these changes were shared in part across MDD, BD, and SCZ.

Nearly all cell type-specific DE genes shared across disorders are involved in aspects of the formation and maintenance of neuronal circuits. For example, *NRXN3* (coding for neurexin 3) was downregulated in PVALB cells in BD (log_2_ fold change [logFC] = −0.38, FDR = 0.03) and MDD (logFC = −0.56, FDR = 9.0 × 10^−12^), and downregulated in VIP cells in SCZ (logFC = 0.33, FDR = 1.4 × 10^−4^) and MDD (logFC = −0.23, FDR = 3.6 × 10^−10^). *NRXN3* is part of a family of presynaptic single-pass transmembrane proteins that act as synaptic organizers, and genomic alterations in *NRXN3* have been identified in a number of neuropsychiatric disorders, including autism spectrum disorders (ASD), schizophrenia, intellectual disability (ID), and addiction (53). *CNTNAP2* (coding for Contactin associated Protein 2) also belongs to the neurexin family and is required for the organization of myelinated axons, and was downregulated in VIP cells in SCZ (logFC = −0.42, FDR = 5.1 × 10^−4^) and MDD (logFC = −0.27, FDR = 0.18). Heterozygous missense and loss-of-function (LOF) *CNTNAP2* mutations cause several neurodevelopmental disorders, including ASD and ID (54), likely through the disruption of synaptic transmission, neuronal connectivity, and network-level activity as identified in LOF mouse models (55,56). As another example, *ERBB4* (coding for Erb-B2 Receptor Tyrosine Kinase 4) was downregulated in VIP cells in SCZ (logFC = −0.56, FDR = 5.1 × 10^−12^) and MDD (logFC = −0.36, FDR = 0.014), and *NRG3* (coding for Neuregulin 3) was downregulated in SST cells in BD (logFC = −0.20, FDR = 5.0 × 10^−20^) and MDD (logFC = −0.23, FDR = 1.2 × 10^−15^). *ERBB4* codes for different isoforms of the receptor tyrosine-protein kinase, which binds its ligand, *NRG3*, a trophic factor. Together, *ERBB4* and *NRG3* play a critical role in the establishment and function of neuronal circuits (57), and their disruption increases susceptibility to SCZ and BD (58). Other examples of affected genes regulating neuronal circuits are *NPAS3, FGF14* and *AUTS2.* Genes regulating neuronal signaling, including *DLG2, GRIK2 and KCNIP4*, were also DE across disorders. See **Supplemental Figure 5** for the full list of shared genes.

We compared our cell type-specific patterns of differential expression to those from a prior meta-analysis of microarray and RNA-seq gene expression in bulk neocortical samples (45) (Figure 1E). A total of 43 cell type-specific DE genes from our analyses were convergent (same logFC sign, FDR < 0.20) with DE changes in bulk tissue (p = 7.08 × 10^−57^, hypergeometric test), with the most overlaps in PVALB cells in SCZ (N = 13). As one example, bulk RNA-seq identified the upregulation of *NPAS3* in SCZ (logFC = 0.14, p = 5.5 × 10^−4^), a change that was uniquely observed in SST cells in our analyses (logFC = 0.36, p = 7.4 × 10^−6^). *NPAS3* plays a broad role in neurogenesis, and mutations in this gene have been associated with SCZ and neurodevelopmental disorders (59,60). LCM-seq cell-specific disease signatures were also moderately consistent with those surveyed in recent human snRNA-seq datasets from MDD and SCZ (30,34), as assessed by rank-rank hypergeometric overlap analysis (**Supplemental Figure 4**, see Methods).

### Neuronal subtypes exhibit distinct and coordinated dysregulation of biological processes across psychiatric disorders

To characterize the biological effects of transcriptomic dysregulation in neuronal subtypes, we performed differential pathway activity analysis. We focused on 1881 pathways representing eight broad biological themes, including bioenergetics, neuron structure and function, and inflammation, known *a priori* to be relevant to psychiatric disorders. For each cell type and pathway, we defined an “activity score” summarizing the collective expression levels of genes in the pathway for that cell type, and diagnosis-specific differences were calculated using multivariate linear models (see Methods).

Figure 2A presents all significantly altered pathways (N = 276, p < 0.01), organized by biological theme. The greatest number of perturbed pathways were in VIP (N = 115), SST (N = 93), and L5/6 PYR (N = 100) cells in SCZ. Although alterations varied between cell types, on average, 11.3% of significant pathway changes within an individual cell type were also significantly altered in the same direction in another disorder. The highest overlap of significant pathways between disorders was observed between MDD and SCZ in VIP cells (31.6%, p = 0.026, hypergeometric test).

**Figure 2.**
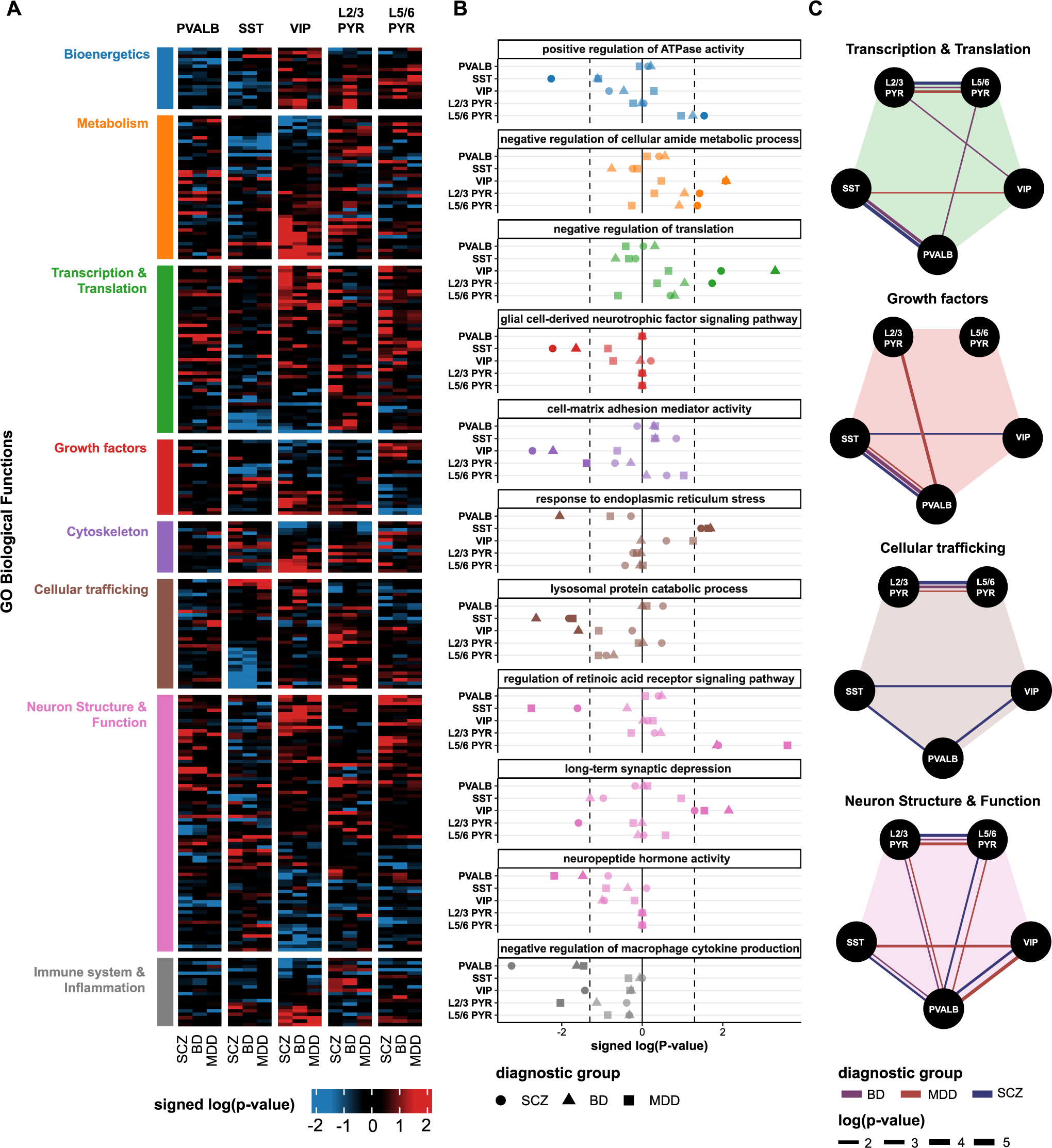
Cortical neuron subtypes exhibit disruption of biological pathways across psychiatric disorders. **(A, B)** Differential pathway activity analysis results. **A)** Heatmap of disease-associated GO biological pathways (rows) altered in various cell type-disorder contrasts (columns), grouped by biological theme. Red and blue colors represent the significance (signed −log10 p-value) of up- and down-regulated pathway activities, respectively. See Supplemental Table 2 for the full list of pathways. **B)** Differential pathway activity (x-axis, signed −log10 p-value) of select biological processes in each cell type (rows) and disorder (shape) (dashed line = p < 0.05). **C)** Similarity of biological pathway alterations between neuronal subtypes across disorders, indicated by the significance (line width, −log10 p-value) of the Pearson correlation of genes in various biological themes (same as A,B) between cell types and across disorders (edge color). All correlations are positive. See Supplemental Figure 6 for all eight biological themes.

Figure 2B highlights cell type-specific differential activities for pathways that reflect known and novel aspects of cellular pathology in psychiatric disorders. For SST cells, this included the downregulation of genes involved in ATPase activity in SCZ (p = 5.5 × 10^−3^), glial-cell derived neurotrophic factor receptor signaling in SCZ and BD (p = 6.0 × 10^−3^, p = 2.3 × 10^−3^), lysosomal protein catabolic process (p = 2.3 × 10^−3^), and retinoic acid receptor signaling in SCZ and MDD (p = 0.025, p = 1.8 × 10^−3^). Notably, there was an upregulation of genes involved in response to endoplasmic reticulum (ER) stress in SST cells for all three disorders (SCZ: p = 0.034, BD: p = 0.02, MDD: p = 0.023), corresponding to cell type-specific findings in a chronic psychosocial stress model of depression in rodents (25). For VIP cells, there was increased negative regulation of genes involved in cellular amide metabolism in BD and MDD (p = 8.2 × 10^−3^, p = 8.5 × 10^−3^), translation in SCZ and BD (p = 0.011, p = 5.0 × 10^−4^), and cell-matrix adhesion mediator activity in SCZ and BD (p = 1.9 10^−3^, p = 6.1 × 10^−3^). For PVALB cells, there was an upregulation of genes involved in long-term synaptic depression in BD and MDD (p = 7.1 × 10^−3^, p = 0.029) and a downregulation of genes involved in oxidoreductase activity in BD and MDD (p = 4.8 × 10^−4^, p = 5.6 × 10^−3^). A full list organized per cell type, themes, and biological pathways is available in **Supplemental Table 2**.

We next assessed the degree of shared alterations in biological pathways between cortical microcircuit neuronal cell types within each disorder. For each of the eight biological themes, we tabulated the set of genes found in at least one pathway associated with that theme. Then, for each theme and disorder, we calculated the Pearson correlation of DE *t*-statistics for these genes between each pair of neuronal subtypes. There were no significant negative correlations; only coordinated changes were observed (Figure 2C). Several findings emerge from this analysis. First, L2/3 PYR–L5/6 PYR and SST–PV cell pairs showed frequent coordinated changes for 6/8 and 5/8 biological themes, respectively (**Supplemental Figure 6**). Second, genes related to transcription and translation, cellular trafficking, and neuron structure and function were correlated between L2/3 PYR and L5/6 PYR cells for all three disorders, as were growth factor-related genes between SST and PVALB cells. Third, genes in neuron structure and function pathways had the most pairwise correlations between cell types, indicating coordinated changes at the microcircuit level. In contrast, other pairs of cells showed much fewer correlated gene changes across disorders, such as VIP–PV, VIP–PYRs, SST–PYRs or SST–PVALB.

### Cell type-specific differentially expressed genes overlap with known genetic psychiatric risk genes

We asked whether cell type-specific DE genes overlapped with risk genes identified by genome-wide association studies (GWAS) of psychiatric disorders. To determine candidate causal genes that mediate the effects of disease-associated genetic variants from recent large-scale GWAS of MDD, BD, and SCZ (51,52,61) we utilized PsyOPS and PoPS, two methods that prioritize casual genes at GWAS lo ci (62,63) (see Methods). Our analysis revealed 22 candidate causal genes exhibiting DE within one or more neuronal subtypes – almost all of which were expressed in interneurons rather than in PYR cells (Figure 3B). Candidate causal genes were especially enriched for DE effects in PVALB cells in SCZ and MDD (p = 0.015, p = 0.026; permutation test).

**Figure 3.**
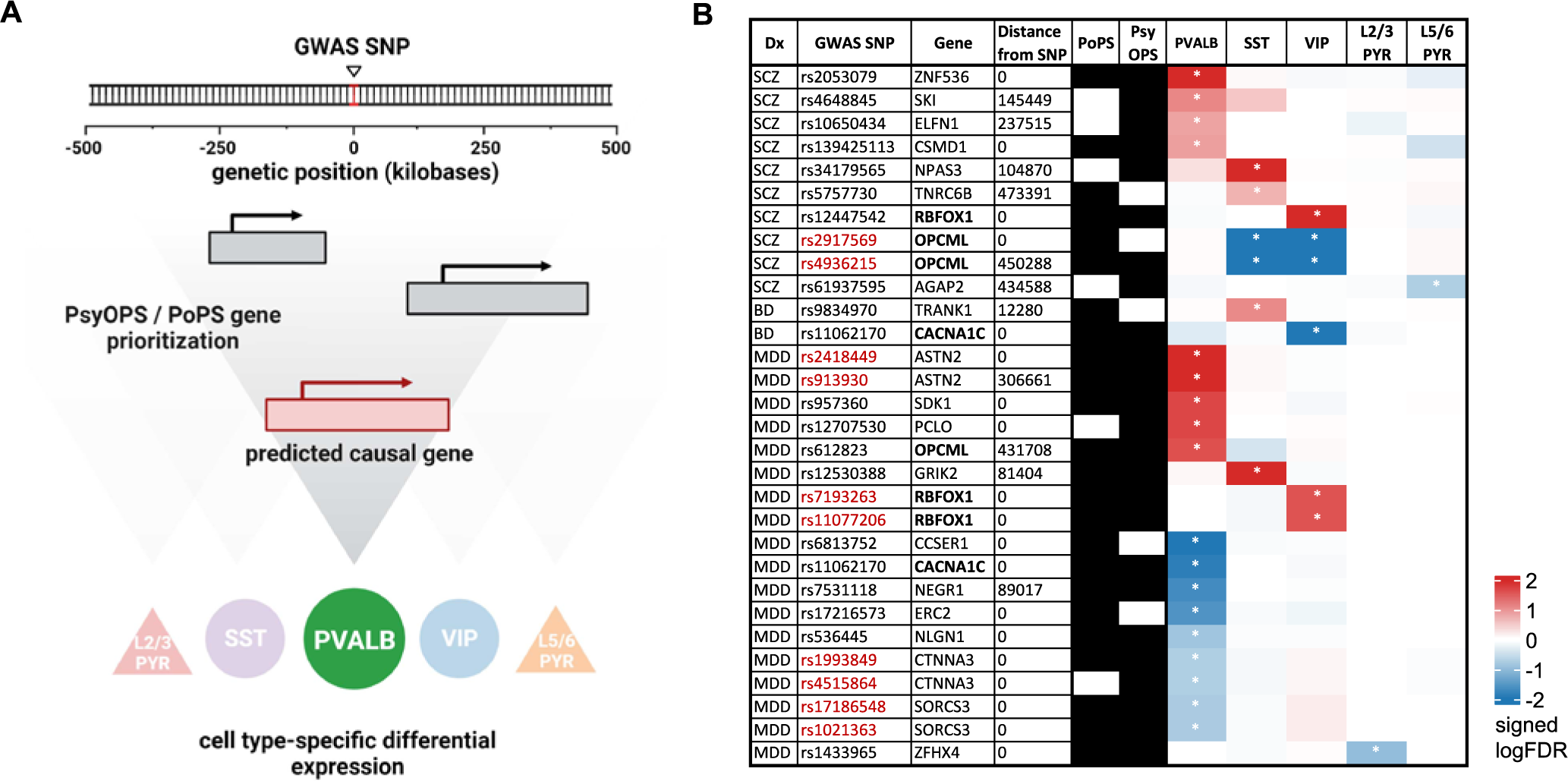
Cell type-specific differentially expressed genes overlap with known genetic risk variants from psychiatric genome-wide association studies. **A)** Overview of approach. Two recent methods, PsyOPS and PoPS (62,63), are used to predict genes that are causally associated with psychiatric GWAS loci, which are then assessed for overlap with LCM-seq differential expression (DE). **B)** 29 unique single-nucleotide polymorphisms (SNPs) associated with MDD, BD or SCZ (diagnosis; Dx) were within 500 kb of a cell type-specific DE gene (FDR < 0.20) from that respective disorder and prioritized as causal by either PsyOPS and/or PoPS. Overlap of SNPs and DE genes was significant for PVALB interneurons in SCZ (p = 0.015) and MDD (p = 0.026, permutation test). Multiple SNPs can be prioritized to the same gene (red text), and three genes (*RBFOX1*, *OPCML*, *CACNA1C*) were shared between two disorders (bold text).

For example, we found that the lead variant (i.e., the most significant GWAS variant within the link disequilibrium block) rs12447542, associated with SCZ, and rs7193263 and rs11077206, two independent lead variants associated with MDD, were predicted by PoPS and PsyOPS to casually effect the *RBFOX1* gene (Figure 3B). *RBFOX1* codes for a neuron-specific RNA-binding protein and splicing factor, and the *RBFOX1* locus is highly pleiotropic, being associated with many psychiatric diseases (64,65). We found that *RBFOX1* is upregulated uniquely in VIP cells – and no other cell type – across all three disorders (SCZ logFC = 0.41, FDR = 4.4 × 10^−7^; BD logFC = 0.53, FDR = 2.97 × 10^−6^; MDD logFC = 0.22, FDR = 0.025), supporting a role for VIP cells as a key mediator of *RBFOX1’s* effects on diverse psychiatric disorders. However, it is worth noting that *RBFOX1* has been shown through experimental studies to also regulate splicing and neuronal function in other types of interneurons, including PV and SST (66,67). Similarly, rs2053079, associated with SCZ, is predicted to have *ZNF563* as the causal gene. *ZNF563* is a zinc finger nuclease that regulates neuronal differentiation in the developing brain (68), and we observe that this risk gene is specifically upregulated in PVALB cells for SCZ (logFC = 0.61, FDR = 4.45 × 10^−3^). As a third example, rs7758630, associated with MDD, is predicted to have *GRIK2* as the causal gene. *GRIK2*, which encodes a subunit of the kainate glutamate receptor, was upregulated in SST cells in SCZ (logFC = 0.61, FDR = 6.4 × 10^−23^), BD (logFC = 0.61, FDR = 2.8 × 10^−23^), and MDD (logFC = 0.32, FDR = 1.4 × 10^−12^) (**Fig 1F**).

## Discussion

We present the first dataset of gene expression in key neuronal cell types forming neocortical microcircuits (PVALB, SST, VIP, PYR) across multiple psychiatric disorders (MDD, BD, SCZ). Alterations in cell types that form cortical microcircuits, the fundamental units of information processing, can lead to significant circuit and network changes, resulting in symptoms and clinical manifestations of psychiatric disorders (26). Previous studies have reported transcriptional changes in individual cell types (35–38), but not at the microcircuit level. Here we performed LCM-seq in postmortem subgenual anterior cingulate cortex samples from 76 subjects, evenly split between MDD, BD and SCZ cases, and healthy controls. We found that disease-associated DE genes were predominantly enriched in interneuron subtypes over PYRs and partly shared across disorders, particularly those related to neuronal circuit formation and maintenance. While biological pathway changes mostly varied between cell types, some had shared alterations among specific cell pairs, biological themes, and disorders. Notably, genes involved in neuron structure and function exhibited coordinated changes across the microcircuit. Finally, DE genes aligned with known risk variants from psychiatric genome-wide studies, showing a cell type-specific link between genetic and transcriptomic risk. These findings indicate both distinct biological alterations and a consistent set of structural and functional changes in neocortical microcircuits across major psychiatric disorders.

In our study, the LCM-seq method addresses the numerical disparity between neuronal subtypes; approximately 80% of cortical neurons are PYRs. (69). We profiled 130 cells of each subtype per individual, ensuring equal representation for rarer interneuron subtypes. Our DE analysis indicates that interneurons, especially PVALB, followed by SST and VIP, appear most altered in psychiatric disorders. This is in contrast to the preponderance of DE genes among excitatory neurons (relative to inhibitory) reported by recent single-nucleus studies of psychiatric disorders (29–34), which might be influenced by higher sampling of excitatory neurons (**Supplementary Figure 7**). In our study, few DE genes were found overall in PYR cells. We note, however, that we may have been under-powered to detect DE in excitatory neurons, as samples were pooled from a relatively broad array of excitatory neuron subtypes. Still, in the present study and with the LCM-seq approach, inhibitory neurons seem transcriptionally more altered in psychiatric disorders than excitatory neurons. Notably, PVALB cells appear particularly affected, aligning with prior studies and theories on psychiatric disorder pathophysiology (23,37,70,71)

We found a notable overlap in DE genes and impacted biological functions across cell types and psychiatric disorders. While DE genes were predominantly cell type-specific, 9.2% changed in the same direction in at least two disorders – far more than expected by chance (p = 2.6 × 10^−33^). Almost all shared DE genes are important for neuronal circuit formation or maintenance, especially in neurogenesis (*AUST2, CNTNAP2, ERBB4, FGF14, LSAMP, NRG3, NPAS3, RBFOX1, ZNF536*), neuronal adhesion (*CLIP3, DSCAML1, KAZN, MAGI2, NRXN3, NXPH1, TIAM1*), and synaptic transmission (*CADPS, FRMPD4, GRIK2, KCNIP4*). We also noticed transdiagnostic disruptions in biological processes, including glial neurotrophic factor signaling, response to endoplasmic reticulum stress, dendrite development, and regulation of translation. The sharing of differential expression signatures and affected biological pathways parallels the known sharing of bulk-tissue differential expression signatures (45), brain imaging signatures (72–76), and genetic risk factors (77,78) across these disorders. We found a number of DE genes overlapping with risk genes from psychiatric GWAS (Figure 3). Genetic risk prominently manifested in PVALB cells in SCZ and MDD, less so in SST and VIP cells in SCZ, and scarcely in PYRs in the three studied disorders.

We further examined coordinated changes in biological processes across cell types, reflecting potential (mal)adaptive states of cortical microcircuits in psychiatric disorders. Using genes that define biological pathways and themes, our analyses revealed that PVALB – SST and L2/3 PYR – L5/6 PYR cell type pairs experience coordinated shifts at the pathway level. The taxonomic relationship between cell types could explain this observation; for example, SST and PVALB cells originate from the medial ganglionic eminence (28) and L2/3 and L5/6 PYR cells are related excitatory cell types. We reason that cell types with overlapping molecular programs may be impacted similarly by psychiatric disorders. Relatedly, such cell types may adopt similar functional adaptations. For example, PVALB and SST cells modulate the excitatory output from and input onto PYR cells, therefore these cell types might coordinate changes to maintain PYR cell activity. Our work using similar methods applied to rodent stress models support this, showing molecular changes indicative of reduced SST cell activity and increased PVALB input to PYR cells (25). Similarly, electrophysiological studies also found reduced inhibitory input to PYR cells after periods of extended stress (79). Though human disease and rodent stress studies differ in their time frames, both demonstrate coordinated dysregulation of similar biological pathways that converge on neuron structure and function. These findings suggest long-term (mal)adaptive alterations of the cortical microcircuit in PVALB and SST cells resulting in changes to excitatory-inhibitory balance in PYR cells.

This study is not without limitations. First, while sex-specific differences are known to exist for these disorders (30,80), we focused on resolving cell-type and disorder differences rather than increasing the number of subjects that would be required for a sex-stratified analysis. The potential contribution of sex and other biological (age, race), clinical (mode of death, medication), technical and other (tobacco use) covariates were assessed and presented in **Supplemental Figure 2**. Second, we cannot resolve sub-subtypes of our five neuronal populations, which may be selectively vulnerable in the context of brain disease (29–34,81). Third, profiling only 130 cells per neuronal subtype limits power, compared to single-nucleus datasets with hundreds of thousands to millions of cells, albeit with strong bias towards common cell types (Supplementary Figure 7). However, our LCM-seq approach is compensated by a deeper sequencing rate per cell type (see Methods). Despite the relatively large technical differences between our study and related studies using more recent technologies like snRNA-seq, we were heartened that at the level of the full transcriptome, the patterns of DE were mostly concordant across studies (**Supplementary Figure 3, 4**).

In sum, we present the first dataset of neuronal subtype-specific gene expression changes across major psychiatric disorders and neuronal subtypes forming canonical cortical microcircuits. We reveal partially shared and concerted biological (mal)adaptive changes across microcircuit cell types and across disorders, suggesting changes that may be constrained by the conserved function of cortical microcircuits, despite additional psychiatric disorder-specific pathologies. Since changes in any cortical microcircuit cell type affect basic aspects of cortical information processing (82,83), they may contribute to the cognitive deficits that are similarly observed across all the investigated psychiatric disorders. Our study lays the groundwork for larger-scale studies investigating transdiagnostic changes across psychiatric disorders in specific brain cell types.

## Data Availability

All data produced in the present study are available upon reasonable request to the authors.

## Acknowledgements and Disclosures

This work was supported by Canadian Institute of Health Research Project Grant No. 153175 (to ES), CAMH Foundation Discovery Fund (to SJT), CAMH Foundation Krembil Startup Fund (to SJT), Ontario Graduate Scholarship (to KA and DJN), and Canadian Open Neuroscience Platform Initiative (to DJN).

Conceptualization, KA, DJN, SJT, and ES; Formal analysis, KA, DFN, and MCD; Investigation, DFN and HO; Resources, DAL and ES; Writing – original draft, KA, DJN, MW, SJT, and ES; Writing – Review & Editing; KA, DJN, MCD, DAL, MW, SJT, and ES; Visualization, KA; Supervision, SJT and ES.

ES is co-founder, CEO and CSO of Damona Pharmaceutical Inc., a biotech company developing GABAergic molecules for the treatment of cognitive dysfunctions in depression. The remaining authors declare that the research was conducted in the absence of any commercial or financial relationships that could be construed as a potential conflict of interest.

## Supplemental figures

**Supplemental Figure 1.**
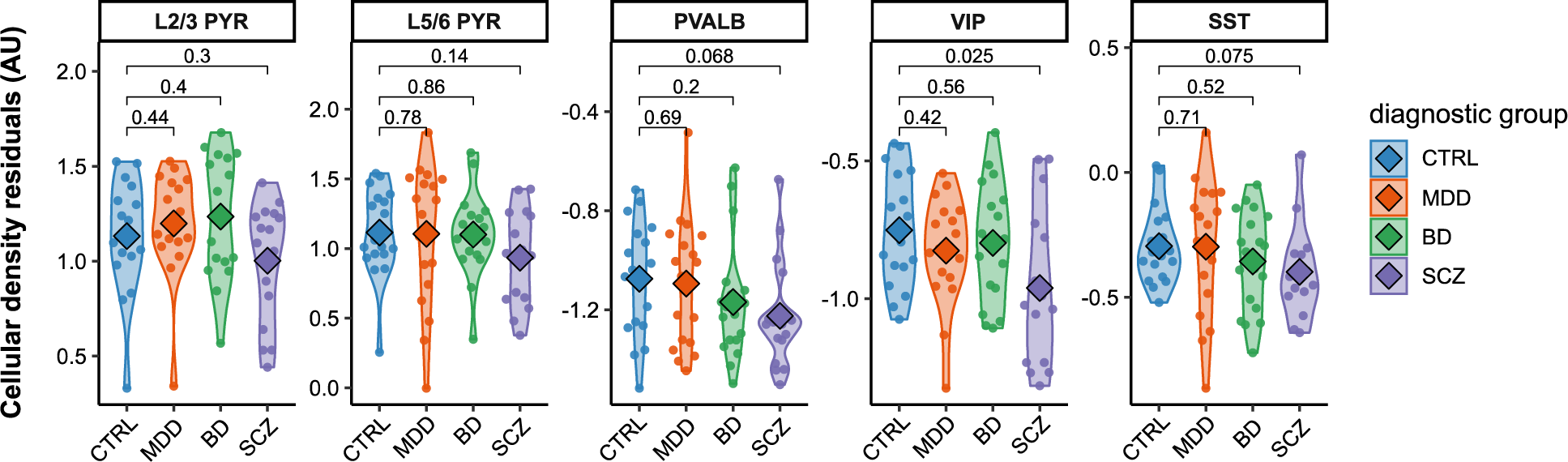
Stereological cell densities of cortical neuron subtypes across disorders. We performed double-label fluorescent in situ hybridization (FISH) on sgACC sections, targeting Slc17a7 (a marker of pyramidal cells) + Sst, or Pvalb + Vip, and counterstained with DAPI to determine changes in cell type density. We adopted a stereological approach to randomly sample 20 sites of the sgACC for imaging. We distinguished L2/3 and L5/6 pyramidal (PYR) cells through the selection of sites superficial and deep to a reference line midway between the pial surface and white matter. We obtained 12 Z-stacks of images that represented transverse depth into tissue sections at a step size of 0.25 μm, which were collapsed into a composite image, normalizing intensities across all samples. We manually quantified cells containing 10 or more cell-marker grains and overlapping with DAPI. We adjusted cell type densities for age, sex, and PMI, obtaining cellular density residuals (arbitrary units, AU), before making case-control comparisons with Wilcoxon signed-rank tests (p-values show).

**Supplemental Figure 2.**
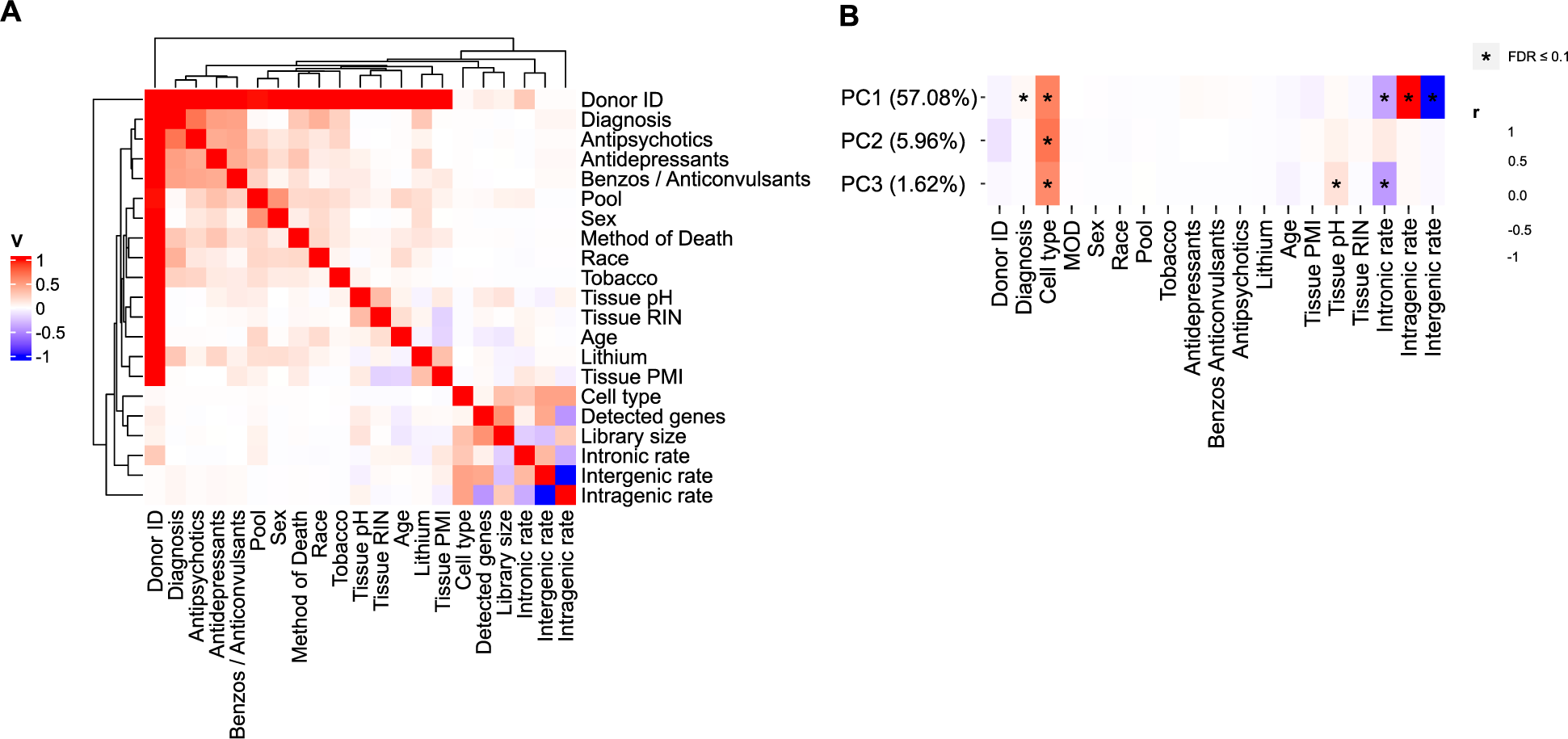
Covariate exploration. We examined 18 biological and technical covariates. Biological covariates comprised sex, age, race, method of death (MOD), tobacco use, medication use (antipsychotics, antidepressants, lithium, benzodiazepines and/or anticonvulsants), while technical covariates comprised pH, RIN, PMI, pool (batch), number of detected genes, library size, intronic rate, intergenic rate, and intragenic rate. A) We calculated Cramér’s V statistics, a measure of correlation between categorical variables, for each pair of biological and technical covariates. B) We also assessed the Pearson correlations between each covariate and each of the top three principal components (PCs) of normalized gene expression. Significant correlations (FDR ≤ 0.10) are starred. Bracketed numbers after each PC denote the percent of variance in gene expression the PC explains. PMI = postmortem interval, RIN = RNA integrity number (measured in tissue homogenates).

**Supplemental Figure 3.**
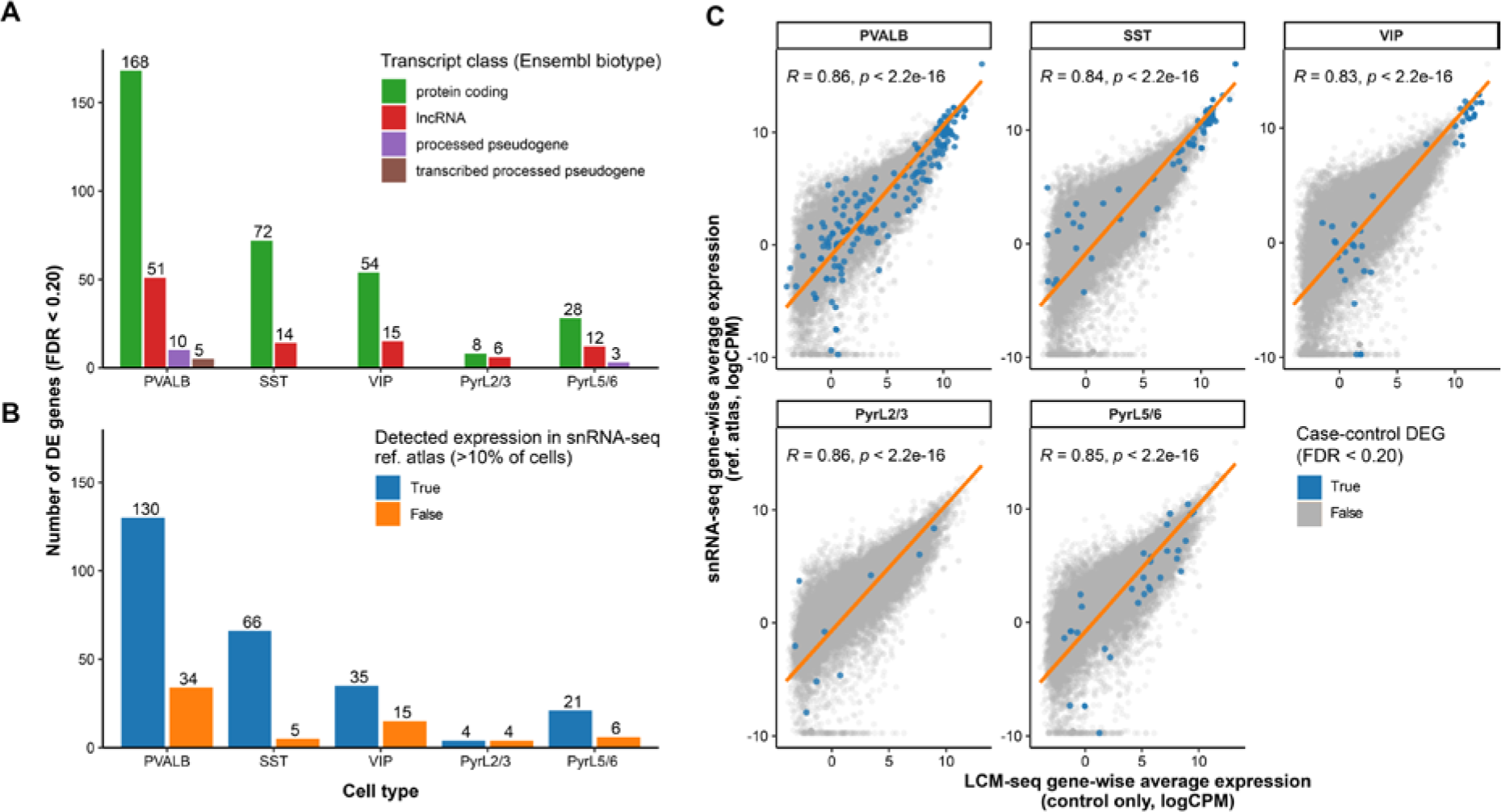
Examining differentially expressed genes. A) We annotated differentially expressed (DE) genes in each cell type with their transcript class. B) We evaluated the specificity of cell type-specific DE genes using an external single nucleus RNA-seq dataset of human medial temporal gyrus from the Allen Institute (46). First, we mapped each of the five LCM-seq neuronal subtypes to its closest snRNA-seq ‘subclass’ based on maximal expression of subclass-specific marker genes. We use blue to denote DE genes that are cell type-specific, where at least 10% of cells of the respective snRNA-seq subclass expressed that gene. C) We calculated the gene-wise correlation of expression levels between LCM-seq controls and the snRNA-seq reference to assess concordance between these sequencing methods. We reported Pearson correlations and p-values. We denoted case-control DE genes in blue, and the orange line was y = x.

**Supplemental Figure 4.**
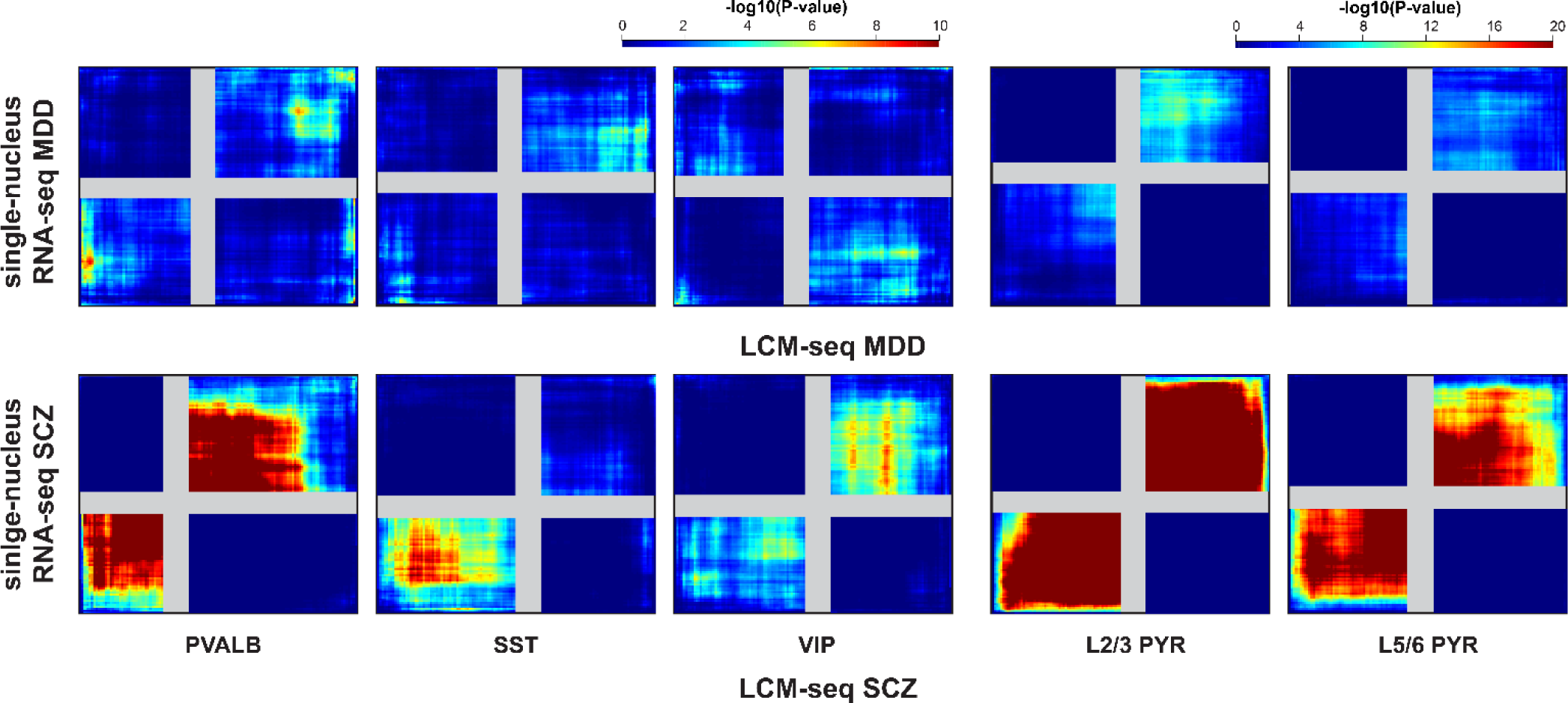
Correspondence between LCM-seq and snRNA-seq differential expression in major depressive disorder and schizophrenia. We further compared the concordance between LCM-seq gene signatures and recent snRNA-seq datasets of MDD (top row) and SCZ (bottom row) (30,34). To assess the overlap of case-control gene signatures between LCM-seq and snRNA-seq in specific cell types, we employed the rank-rank hypergeometric overlap test (RRHO2) (47). Genes were ranked according to their p-value and effect size direction, allowing us to identify significantly overlapping genes across a continuous range of significance levels. Hot regions in the top-right and bottom-left quadrants indicate concordant overlap in the genes between LCM-seq (x-axis) and snRNA-seq (y-axis).

**Supplemental Figure 5.**
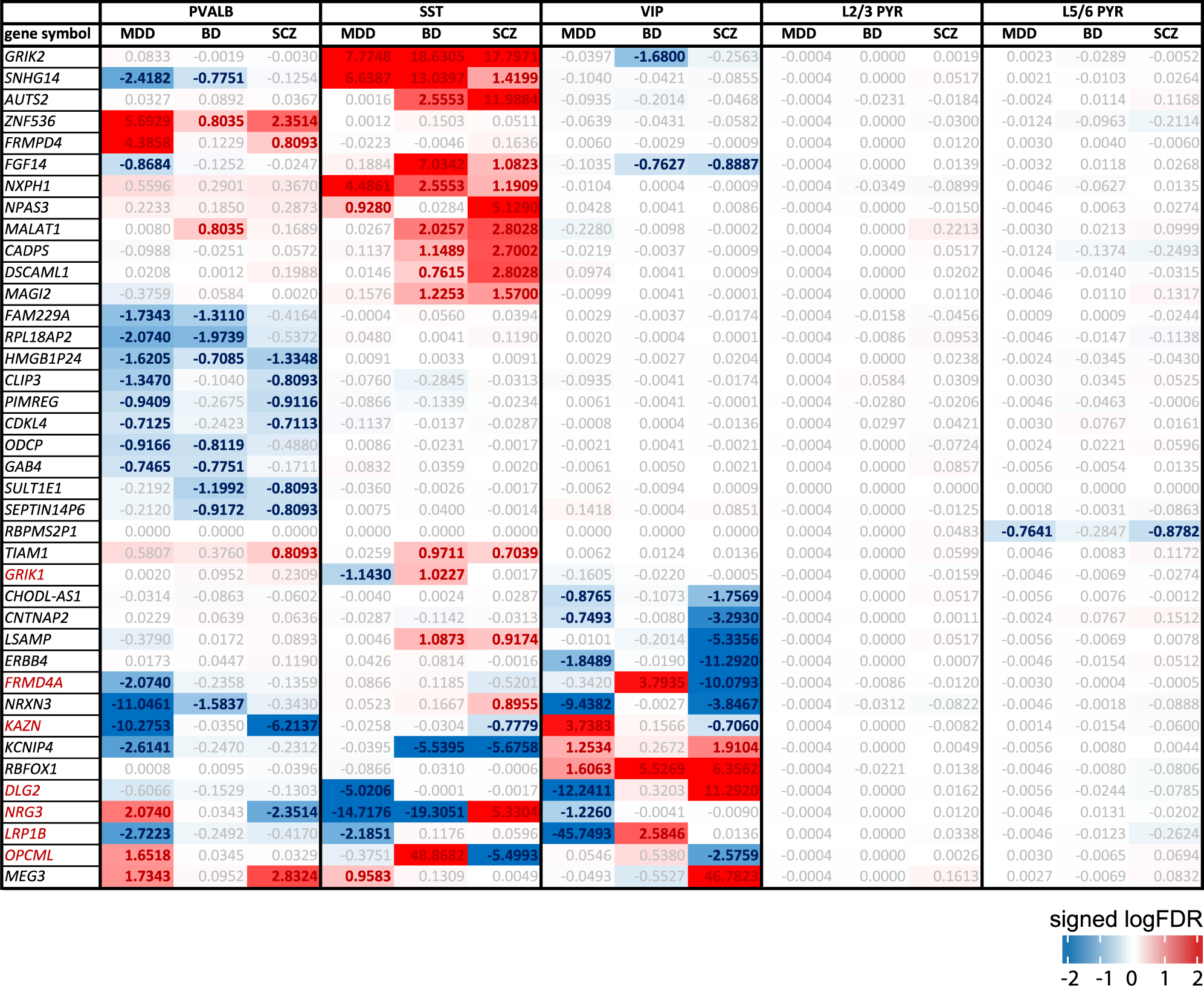
Cell type-specific differentially expressed genes shared across disorders. We identified 39 unique genes that are differentially expressed (FDR < 0.20) in the same cell type for multiple disorders. 34 genes show consistent changes (same logFC), while 7 show opposite changes (opposite logFC, names in red).

**Supplemental Figure 6.**
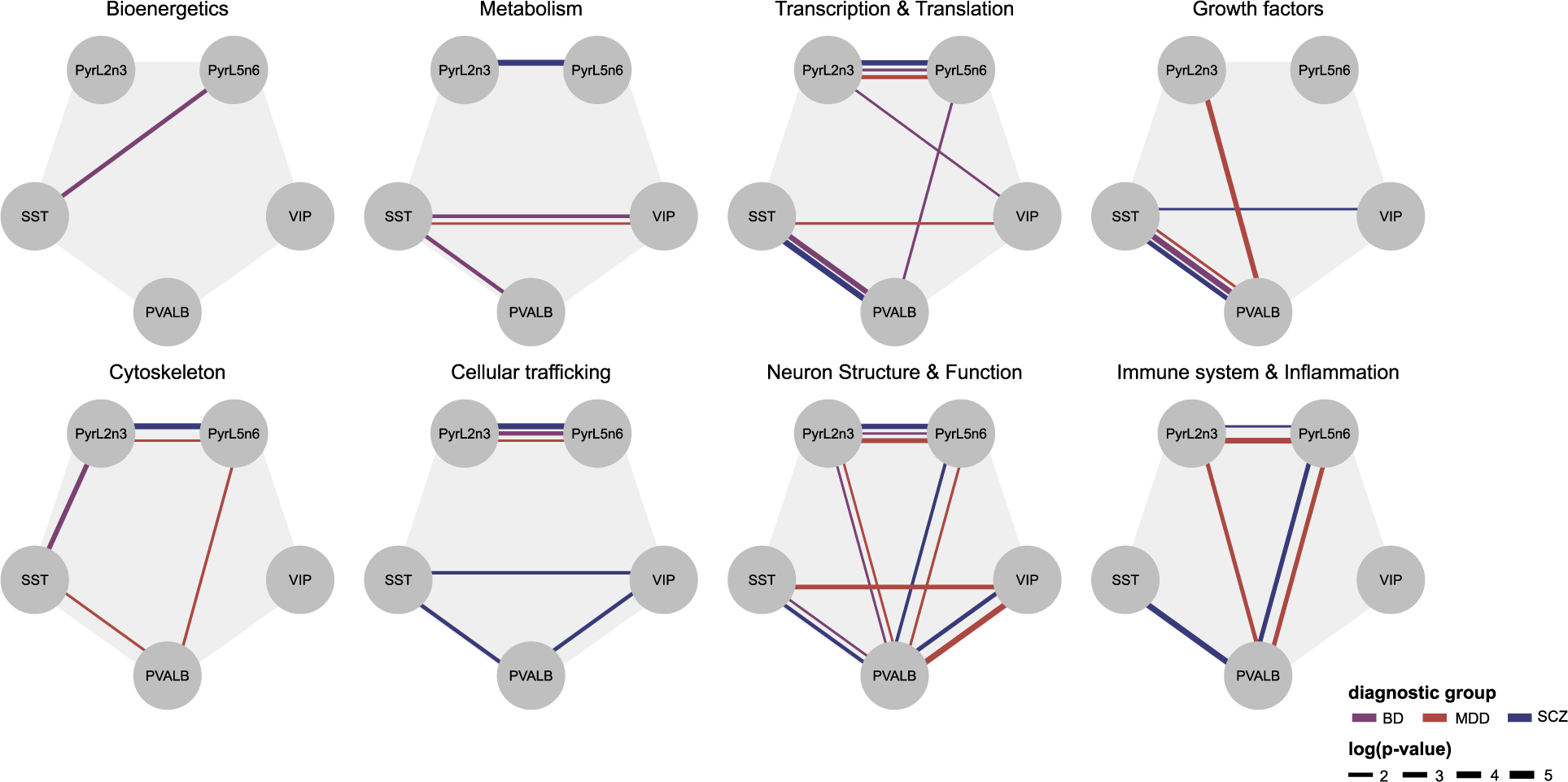
Similarity of biological pathway alterations between neuronal subtypes across disorders. We analyzed the similarity of biological pathway alterations between neuronal subtypes across disorders, indicated by the significance (line width, −log10 p-value) of the Pearson correlation of genes for all biological themes, between cell type pairs and across disorders (edge color).

**Supplemental Figure 7.**
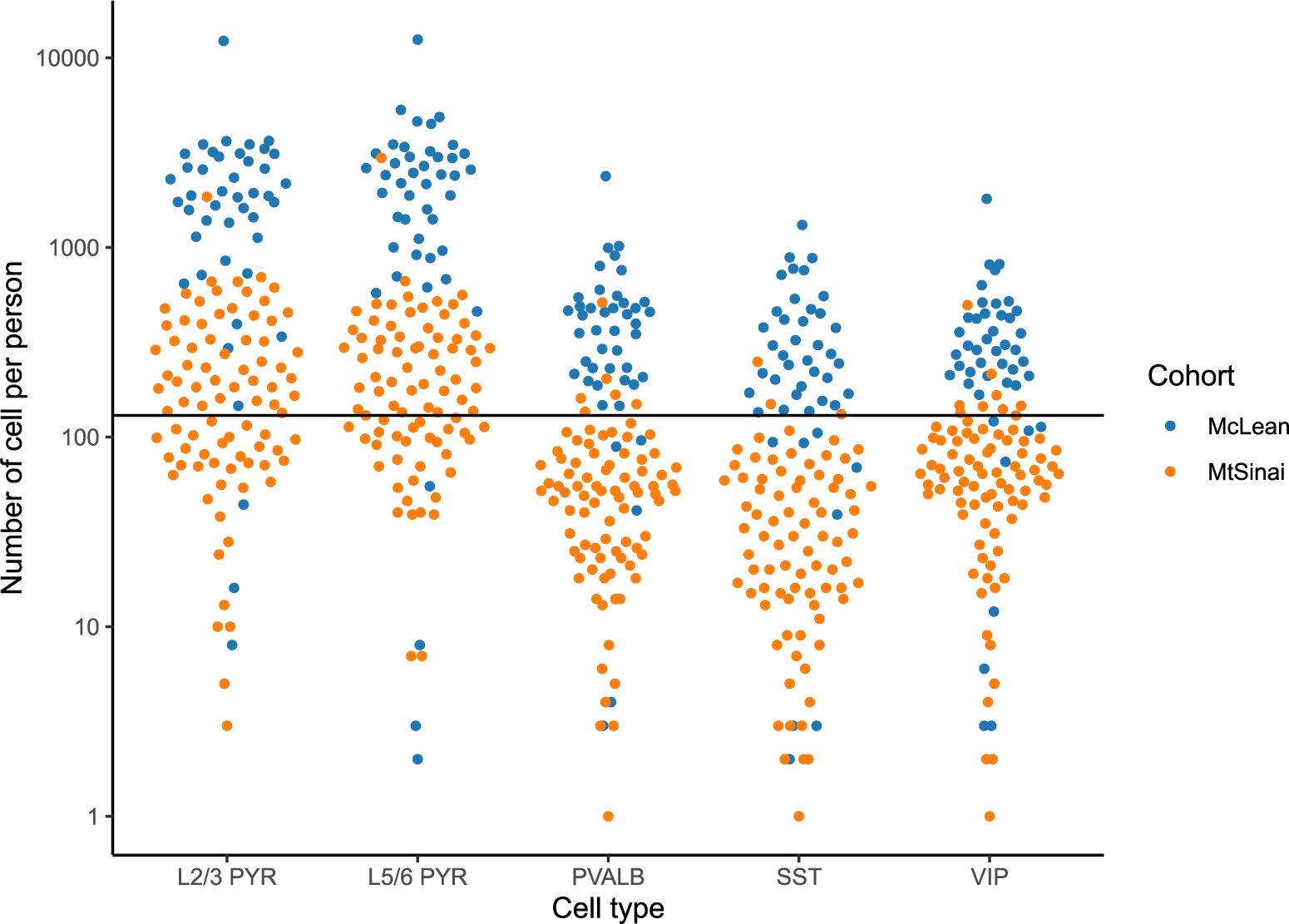
Number of cells sampled per person in the latest snRNA-seq study of schizophrenia (Ruzicka et al. 2022). We show the distribution of the number of cells sampled per person after quality control (y-axis, log scale) in a snRNA-seq study of SCZ in human prefrontal cortex across two independent cohorts (colors), for each neuronal subtype in the current study (x-axis, original cell type-labels from the authors were semantically grouped). The black line represents the 130 cells sampled per person per subtype in the current study.

## Supplemental tables

Supplemental Table 1. *Differential expression analysis results* (Attached)

Supplemental Table 2. *Biological pathway scores across neuronal subtypes and disorders*. (Attached)

